# Is SARS-CoV-2 elimination or mitigation best? Regional and disease characteristics determine the recommended strategy

**DOI:** 10.1101/2024.02.01.24302169

**Authors:** Maria M. Martignoni, Julien Arino, Amy Hurford

## Abstract

Public health responses to the COVID-19 pandemic varied across the world. Some countries (e.g., mainland China, New Zealand, and Taiwan) implemented elimination strategies involving strict travel measures and periods of rigorous nonpharmaceutical interventions (NPIs) in the community, aiming to achieve periods with no disease spread; while others (e.g., many European countries and the United States of America) implemented mitigation strategies involving less strict NPIs for prolonged periods, aiming to limit community spread. Travel measures and community NPIs have high economic and social costs, and there is a need for guidelines that evaluate the appropriateness of an elimination or mitigation strategy in regional contexts. To guide decisions, we identify key criteria and provide indicators and visualizations to help answer each question. Considerations include determining whether disease elimination is: (1) necessary to ensure health care provision; (2) feasible from an epidemiological point of view; and (3) cost effective when considering, in particular, the economic costs of travel measures and treating infections. We discuss our recommendations by considering the regional and economic variability of Canadian provinces and territories, and the epidemiological characteristics of different SARS-CoV-2 variants. While elimination may be a preferable strategy for regions with limited health care capacity, low travel volumes, and few port of entries, mitigation may be more feasible in large urban areas with dense infrastructure, strong economies, and with high connectivity to other regions.

## Introduction

During the COVID-19 pandemic, non-pharmaceutical Interventions (NPIs) greatly reduced the spread of SARS-CoV-2 [39, 15], and include: travel measures, such as self-isolation, quarantine, and testing requirements applying to individuals arriving from other jurisdictions; and community measures, such as physical distancing, gathering size restrictions, and business and school closures, that apply to residents. However, these restrictions substantially reduced economic activity, increased unemployment rates, and undermined social wellbeing [37, 73, 71]. As NPIs have substantial economic and societal costs, it is important to establish criteria to adjust restrictions to control infection spread with minimal cost.

Canada is the second largest country in the world by area, extending from the Pacific to the Atlantic to the Arctic Oceans with ten provinces and three territories widely differing in their geography, population, and economies. Canada is also a federal state, with responsibility for health care divided between the federal government, responsible for: regulation of entry into the national territory; approval of medications and vaccines; the health of First Nations living on reserves, the military, and inmates in the federal prison system, and with the provinces and territories responsible for most of the remaining health care issues, including the implementation of health policies, and entry to, and movement within, their borders. Geographic, economic and jurisdictional differences, as well as differences in health care capacity, have driven broad inter-provincial variation in the type and timing of NPIs implemented to limit viral spread [64, 18, 112], which has in turn led to different COVID-19 epidemics in different Canadian jurisdictions.

Prior to the establishment of the Omicron variant in December 2021, the Atlantic provinces (New Brunswick, Nova Scotia, Prince Edward Island, and Newfoundland and Labrador) and Northern Canada (Nunavut, Yukon, and Northwest Territories) generally implemented a containment strategy [82] that resulted in periods of elimination of community infections [44] consistent with an ‘elimination’ or ‘zero-COVID strategy’ [19, 10, 42] (see Fig. 1a). These provinces and territories have relatively small population sizes [92], and in Newfoundland and Labrador, for example, elimination was achieved through strict border control of few ports of entry (through which travelers from the rest of Canada and abroad could enter the province), contact tracing, testing, and rigorous restrictions to end community transmission when community outbreaks occurred [82]. In contrast, provinces with large urban centers, such as Ontario and Quebec, may have implemented ‘mitigation’ or ‘suppression’ strategies, aiming to flatten the epidemic curve and keep the number of cases below the critical care capacity, with some community transmission [10, 29, 20] (see Fig. 1b). At times during 2020 and 2021, these provinces reported high case counts [47], nearly reaching the critical care capacity in December 2020 and March 2021 [22, 21, 31].

**Fig. 1:**
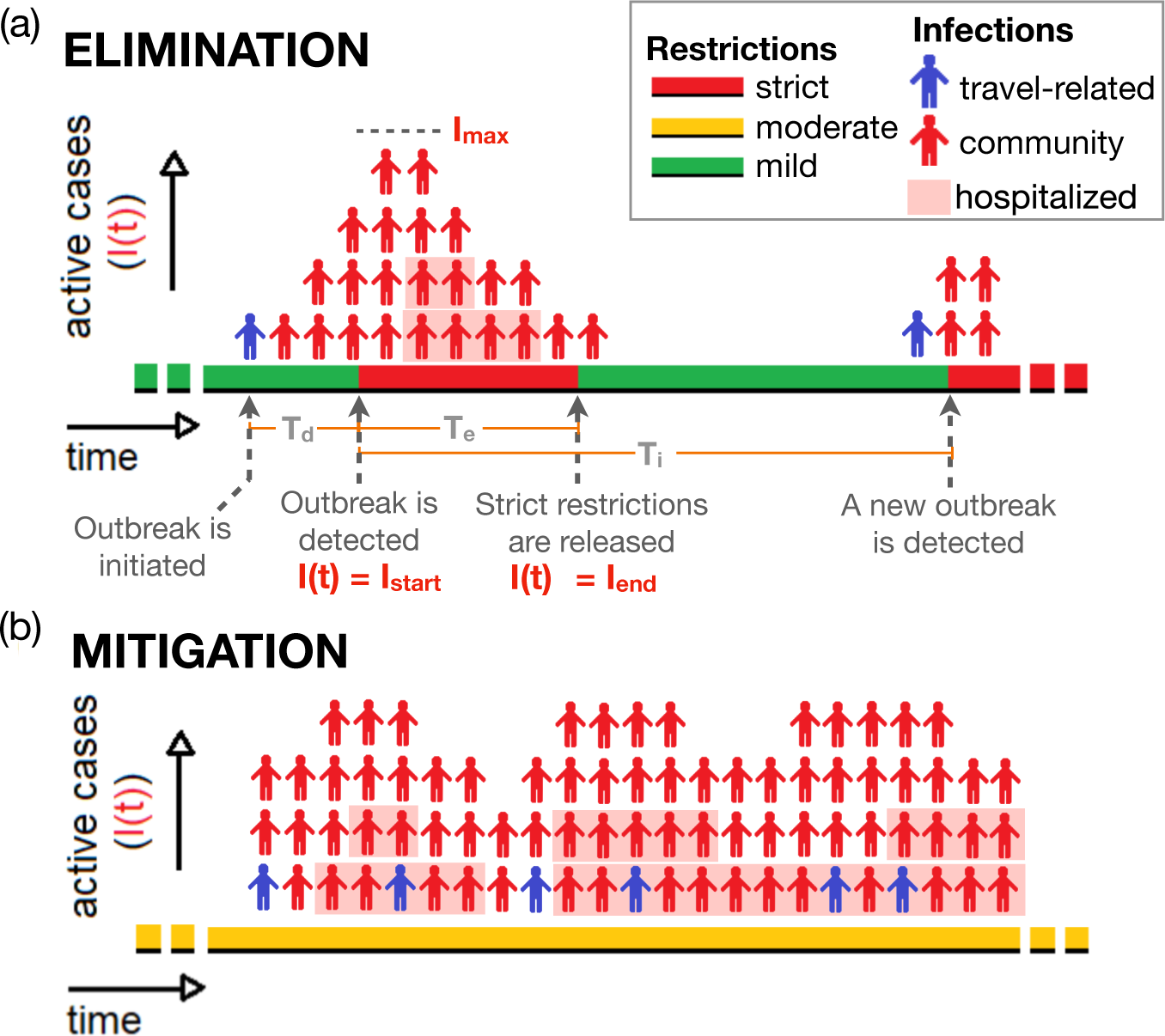
Visual representation of (a) an elimination, and (b) a mitigation strategy and corresponding epidemiological indicators. When an elimination strategy is implemented, a community outbreak initiated by an infected traveler is detected after a time interval *T_d_*. Following outbreak detection, strict restrictions to reduce the number of cases are implemented during the time interval *T_e_* (red regions). Strict restrictions are released when the number of cases drops from *I*_max_ to below a minimum *I*_end_ (green region). The time interval between the time of detection for two consecutive outbreaks is *T_i_*. When a mitigation strategy is implemented, prolonged periods of moderate restrictions are enacted (yellow regions). Red persons correspond to community cases. Blue persons correspond to travel-related cases that infect individuals in the community (i.e., ‘spillover’). For simplicity, travelrelated cases that do not cause community cases are not shown in the figure. Shaded cases correspond to hospitalized cases. The width of a person corresponds to the average duration of an active case and the relationship between incidence, prevalence, and hospital occupancy is investigated in the supplementary information, section A. (Figure adapted from [61]).

Whether an elimination or mitigation strategy is the preferable response to pandemic threats has been matter of debate (e.g., [2, 99, 34, 56, 65, 75]). Early in the COVID-19 pandemic, countries implementing an elimination strategy were praised for obtaining better public health outcomes, economic growth, civil liberties, and general population well-being [74, 10, 9, 40]. However, by late 2021, and especially with more people becoming vaccinated and the spread of the more transmissible Omicron (B.1.1.529) variant of concern, most countries had abandoned the elimination strategy [8, 26, 43], and the societal costs of strict restrictions were increasingly highlighted [13, 68, 106]. These observations indicate that neither elimination, nor mitigation is the indisputable optimal strategy, and that the preferred strategy may vary regionally and change over time.

#### Box 1: Common errors and misconceptions that bias against recommending an elimination strategy

- Multiple studies have shown that travel measures do not have a substantial role when community outbreaks are occurring [7, 105, 101, 16, 24], which may lead to the misconception that travel measures are unimportant. However, in regions with no community cases, travel measures become critical to ensure that mild community restrictions can remain in place for a reasonable period of time.
- Recommendations have considered whether travel measures should be implemented [107, 24], but may have overlooked coordinated implementation of multiple components of a public health response. An elimination strategy might involve travel measures when there are no community cases, an aggressive containment response to community cases when they occur, and a plan to continue the elimination strategy only until the conditions for the implementation of the elimination strategy are no longer met, for example, when the population becomes highly vaccinated [74].
- International guidelines considered as best-practise in several countries [107, 108] are not necessarily applicable to small jurisdictions or remote communities with self-determination of health care (e.g., economically smaller Canadian provinces or Indigenous communities), which may bias against elimination strategy recommendations. While elimination may be unsustainable and economically damaging at the country level, in smaller regions an elimination strategy may provide benefits, for example, helping to protect health system capacity.
- Classic epidemiological models of community spread (e.g. the Susceptible-Infectious-Recovered (SIR) compartmental models, [54]) are frequently used to model infectious diseases, however these models are only applicable to regions implementing an elimination strategy when community outbreaks are occurring. Community spread models are not appropriate to provide reproduction number estimates from data describing travel-related cases, which report infection in individuals that did not acquire infection from the local community, and where the infected traveller may have been in quarantine or self-isolating since arrival. Indeed, the mechanisms that generate new cases when prevalence is low or zero are not the mechanisms that generate new infections in an SIR model, where it is assumed that infected individuals mix amongst susceptible members of the community. This limitation can be overcome, for example, by separately reporting travel-related and community cases, and by estimating the number of outbreaks, and then modelling each community outbreak individually [44, 91, 35].
- Results of stochastic simulations are often reported as means of an ensemble of simulations with community outbreaks starting on different days. The ensemble mean may show no elimination of infection, although elimination does occur for individual simulations. Therefore, the results of stochastic models (including agent-based models) should be carefully reported (e.g. [50]).

Infection severity, health care capacity, the efficiency of case detection, the vaccination status of a population, and the economic and societal costs of NPIs have a fundamental role in determining if an elimination or a mitigation strategy should be preferred. Many such indicators are highlighted in *Guidance for a strategic approach to lifting restrictive public health measures* [81] and other guidance documents by Canadian provincial public health [83], the Public Health Agency of Canada [80], and the World Health Organization [108]. Yet, lacking are quantitative descriptions that specify the relative importance and inter-relatedness of indicators, and how these quantities combine to determine epidemiological quantities that guide decisions.

A further issue that has not previously been highlighted, is that the most relevant epidemiological quantities for regions implementing an elimination strategy are different than for those implementing mitigation or suppression. When a mitigation strategy is implemented, travel-related cases make only a negligible contribution to epidemic dynamics [7, 105, 101], and the infection dynamics are mainly determined by the pathogen spread rate within a community (e.g., by indicators such as the basic or control reproduction numbers of the infection [27, 33, 53]). On the other hand, travel measures, including testing and post-arrival quarantine or self-isolation, may be a more critical component of the combined public health response in regions implementing an elimination strategy. In this case, other forms of assessment such as the evaluation of the efficiency of travel measures, including testing and quarantine policies [116, 97, 79, 96, 6, 91], and the probability of elimination under specific community NPIs (i.e., capacity limits or school and business closures) for hypothetical community outbreak scenarios [78, 14, 41, 35], are key to inform public health responses.

Due to these fundamental differences in the key quantities and modelling approaches used to forecast the epidemiological dynamics of mitigation and elimination, it has been challenging to develop methods that allow for a quantitative comparison of the two strategies for decision-making purposes (see also Box 1, in which we highlight some common errors and misconceptions when deciding whether an elimination strategy should be implemented). Here, we identify key epidemiological and regional characteristics to evaluate whether disease elimination or mitigation is desirable, and we outline criteria to guide this decision. We highlight three main questions that should be answered to determine the circumstances when elimination is a recommended approach (see Fig. 2). Namely, is elimination:

❶ necessary to ensure health care provision?
❷ epidemiologically feasible?
❸ cost effective?

**Fig. 2:**
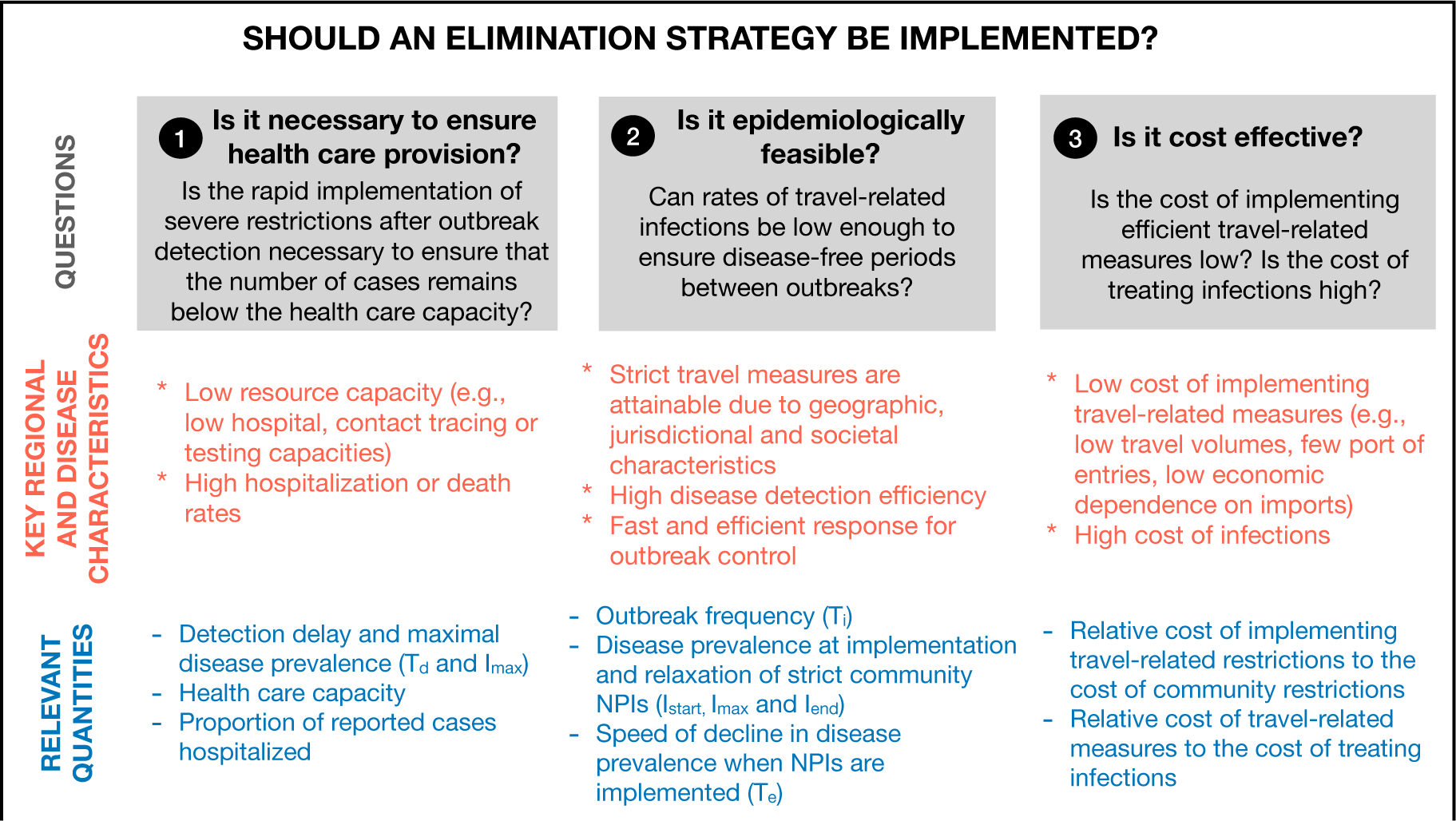
Three possible criteria presented in this manuscript that can be used to determine whether elimination or mitigation strategy is preferable, and key regional and disease characteristics and relevant quantities that should be considered to answer each of the questions. The meaning of the different parameters is shown in Fig. 1.

We discuss each of these questions separately, and consider which role different criteria may play during an unfolding pandemic. Although our discussion mainly focuses on the approach to SARS-CoV-2 of different Canadian provinces and territories, our conclusions have broad implications that apply widely outside the Canadian context.

### ❶ Is elimination necessary to ensure health care provision?

During the pandemic, in many countries a main purpose of NPI implementation was to ensure that the number of severe cases did not rise above hospital or Intense Care Unit (ICU) capacity. High pathogen transmissibility, high rates of asymptomatic cases, and low testing efficiency, are all factors that could cause a community outbreak to go undetected for several days. In regions with low resource capacity (e.g., jurisdictions with low hospital, contact tracing or testing capacities), the number of cases when the outbreak is initially detected may already have the potential to approach, or exceed, available resources for control and health care capacity [12, 102, 45]. This is particularly true of hospital and ICU occupancy limits, because due to the delay between exposure and hospitalization, and because hospital stays are often many days, peak hospital occupancy usually occurs weeks after the implementation of strict community NPIs and can be substantially higher than hospital occupancy at the time of strict NPI implementation. Thus, in these regions the implementation of strict community measures as soon as the outbreak is detected may be necessary to prevent hospital and ICU burden, spurring the implementation of an elimination approach. This choice is accompanied by the implementation of travel measures after the outbreak is eliminated, meaning that the costs and disruptions of strict border control are outweighed by the benefits associated with protecting the capacity of the health care system.

On the other hand, regions with higher resource capacity may be able to detect a new outbreak before the number of hospitalizations reaches the potential to near the regional capacity. In these cases, a disease mitigation approach consisting of moderate community NPIs may be sufficient to ensure hospital capacity limits are not exceeded, and strict travel measures may lead to unnecessary costs and disruptions, that could negatively affect the compliance with other public health measures [115, 30]. Even when health care capacity is high, concerns regarding pathogen variants that are highly transmissible or virulent, and uncertainty in how cases will respond to interventions may justify the precautionary implementation of an elimination approach (or a ‘wait-and-see approach’ [78]) rather than a mitigation approach, in order to delay pathogen spread until more information is available, vaccines or therapy is developed, or response preparedness is enhanced [46, 32, 1, 6].

In the supplementary information, section A, we present an illustrative model to determine hospital occupancy from peak incidence, that can serve as a starting point to quantitatively assess whether there is a risk of exceeding the regional care capacity. The model can also be used to provide estimates for the detection time *T_d_* (see Fig. 1a) between outbreak initiation and detection, and for the corresponding increase in infection prevalence during this time.

Disease severity will also determine whether elimination or mitigation is preferable. The occurrence of severe disease depends on characteristics of a population including the proportion with different ages, co-morbidities, vaccination and immunity statuses, associations between these variables, and where these factors may be heterogeneous within a population [87, 100]. For instance, it may be be reasonable to recommend a SARS-CoV-2 elimination approach for regions whose populations have a high prevalence of co-morbidities, or a low proportion of individuals vaccinated [5, 55]. On the other hand, a lower proportion of cases requiring hospitalization arising from high vaccination rates, or the spread of a less severe variant, may reduce the need for an elimination strategy. The SARS-CoV-2 variant of concern Omicron (B.1.1.529), for example, established and spread in Canada when a large proportion of the population had already received at least two doses of vaccine, reducing the rates of severe disease with respect to other variants (see vertical axis, Fig. 3).

**Fig. 3:**
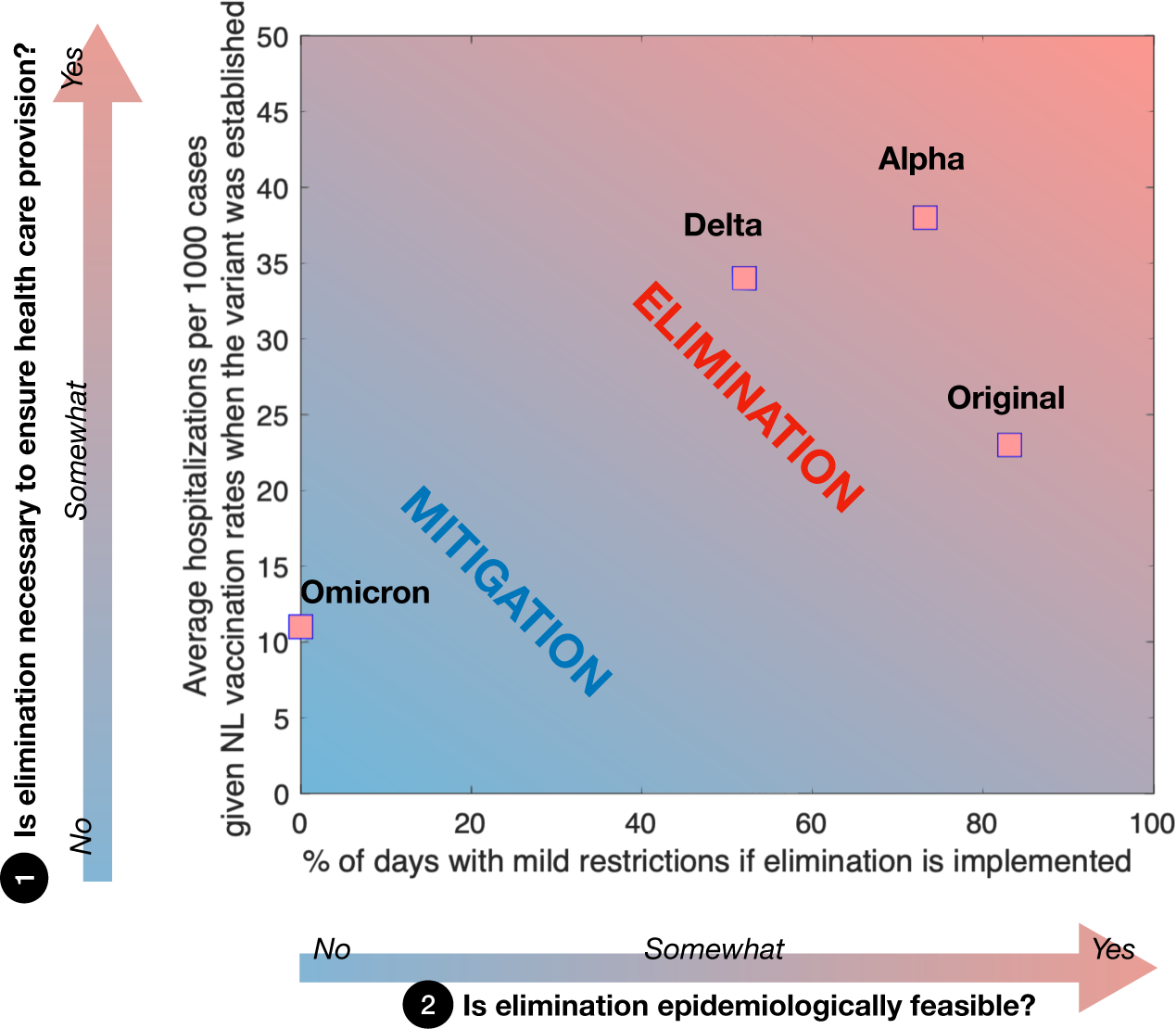
Estimated average hospitalization per 1000 cases considering vaccination rates in the province of Newfoundland and Labrador (NL) at the time each SARS-CoV-2 variant was established (vertical axis) and estimated percent of days with mild NPIs between two consecutive community outbreaks if an elimination strategy is implemented (i.e., 100 *×* (*T_i_ − T_e_*)*/T_i_* for *T_e_ < T_i_* and 0 otherwise, where *T_e_* and *T_i_* are shown on the horizontal axis of Fig. 1a). When high transmissibility does not allow for periods with no community cases between outbreaks, and when the risk of severe disease is relatively low, elimination is no longer feasible, and mitigation is preferred. Estimates used for producing the figure and their derivation are provided in the supplementary information, section B. [Adapted from [61]].

### ❷ Is elimination epidemiologically feasible?

For elimination to be feasible, the duration of the strict restrictions needs to be balanced with a reasonable period when restrictions are relaxed to release the population from the adverse impacts of strict public health measures (i.e., the proportion of green zones to red zones in Fig. 1a needs to be high). This requires both that many weeks elapse between community outbreaks initiated by infected travelers, and that once the outbreak is detected strict community NPIs are implemented sufficiently rapidly to initiate the decrease in infection prevalence.

The probability that a travel-related case initiates a community outbreak depends on travel measures, infection prevalence in neighboring regions, pathogen characteristics, such as its transmissibility, airborne transmission, incubation time or testing efficiency [60, 86, 111], community NPIs and vaccination levels [44]. The time before an outbreak can be considered under control (i.e., when the probability that further community infections may occur is very low) depends on peak incidence and on the effectiveness of the strict NPIs in reducing community transmission. Peak incidence (as discussed in ❶) determines the approximate maximum from which new daily cases must decline, and the effectiveness of strict community NPIs determines the speed of the decline. The speed of the decline depends on pathogen transmissibility, and on the efficiency of contact tracing, testing, and isolation [39, 90]. Thus, local socio-geographic characteristics, pathogen characteristics, and characteristics of the local population can make an elimination strategy more or less feasible from a purely epidemiological point of view, and this feasibility should be continuously reassessed given evolving pathogenic traits and compliance with public health measures.

When a disease is highly transmissible, outbreaks occur often, as prevalence may be higher in other connected regions, and as the virus spreads easily even when restrictions are in place [44], allowing only short or no periods of mild restrictions. For example, it may not have been epidemiologically feasible to eliminate the SARS-CoV-2 Delta variant in Melbourne, Victoria, Australia, even with strict NPIs. On October 21, 2021, Melbourne residents exited an 11 week lockdown because vaccination targets had been met, but at this time daily reported cases in Victoria were 2,232: the second highest that had been reported for any Australian state [49]. In Newfoundland and Labrador there were 10 months between the last reported case associated with the initial community outbreak (original SARS-CoV-2 virus) in March 2020, and the next community outbreak (Alpha variant) that began in February 2021 [82]. The more transmissible and virulent Delta variant was introduced into Newfoundland and Labrador in April 2021, and over the next 8 months several community outbreaks were reported in smaller regions across the province, with much shorter periods between community outbreaks than previously observed [82]. Nonetheless, elimination was achieved (and therefore feasible) in Newfoundland and Labrador for the original, Alpha, and Delta variants, but not for the more transmissible and less virulent Omicron variant.

In Fig. 3 (horizontal axis), we provide estimates of the average percent of days with mild restrictions between two consecutive outbreaks if an elimination strategy is implemented, based on the outbreak frequency experienced in Newfoundland for different SARS-CoV-2 variants of concern. The number of days with mild restrictions depends on the time between outbreaks (i.e., *T_i_*, see horizontal axis of Fig. 1a) and on the time needed to eliminate an outbreak (i.e., *T_e_*, red regions in Fig. 1a), which is calculated assuming an exponential decay in infection prevalence when strict restrictions are in place, as explained in detail in the supplementary information, section B. We calculated that for *T_e_ < T_i_*, the percentage of days with mild restrictions is given by

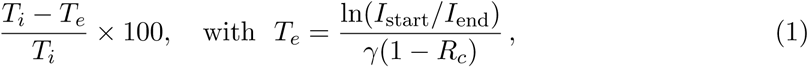

and is equal to zero for *T_e_ ≥ T_i_*. The value of *T_e_* depends on infection prevalence when restrictions are implemented (*I*_start_), on infection prevalence when restrictions are released (*I*_end_), on the infection recovery rate (*γ*), and on the control reproduction number (*R_c_*).

The effectiveness of the same public health measures varies depending on the pathogen or variant considered, making different strategies preferable at different points in time. Additionally, as discussed in ❶, different COVID-19 variants can be characterised by different rates of severe illness, which also depends on vaccination rates. These considerations affect the risk of exceeding hospital capacity (Fig. 3, vertical axis).

Lastly, for an elimination strategy to be feasible it is not only important to achieve fast outbreak detection and implementation of strict community restrictions, but it is also necessary that strict measures are relaxed when they are no longer needed, because during an ongoing pandemic there remains a risk of disease re-introduction, and the population may be asked to comply with strict public health measures once again. The World Health Organization defines an outbreak as over when two incubation periods have passed with no further cases reported (i.e., 28 days for COVID-19 [77]), however, a more precise approach could be to relax measures when there is a high probability that the number of cases in the community is zero [77]), and to consider how quickly the reported cases were isolated. Contact tracing efficiency and population compliance will affect when community NPI relaxation can feasibly occur [63, 17]. Thus, faster reopening may occur in regions characterised by social cohesiveness, such as rural areas where ‘everyone knows everyone’, and where infected people and their contacts are easier to identify and reach [109, 59]. On the other hand, contact tracing might be impractical in larger and more densely populated areas making an elimination strategy more challenging to implement.

### ❸ Is it cost effective?

Discussing costs when it comes to fighting a pandemic threat is challenging, and there are trade-offs to be considered. Minimal public health restrictions may lead to many infections, hospitalizations, and deaths, while strict public health restrictions may lead to economic, social and psychological damages [73, 76]. Considering such costs involves finding a complex balance between medical needs, and social and economic freedom. This said, certain costs, whether social, economic, or medical, are unequivocally larger in some regions relative to others. When deciding if elimination or mitigation is preferable, an important consideration is the trade-off between the economic cost of implementing travel measures, which tend to be higher for a disease elimination strategy; and the costs of treating infections, which tend to be higher for a disease mitigation strategy due to higher number of infections [10, 110].

Travel measures, such as travel declaration forms and testing requirements that are verified at arrival are less costly, and can feasibly be enforced, in regions with few ports of entry. The costs of requiring arriving travelers to quarantine or self-isolate are less in regions with low travel volumes, while the cost of reducing travel-related infections in Canada’s economically larger provinces, i.e., Ontario and Quebec, are substantial due to the large volume of trade occurring across the inter-provincial and international borders [93, 94]. Travel measures may be less costly and more feasible in Atlantic Canada, where there are few ports of entry into most of the four provinces. Furthermore, during the pandemic, international arrivals to Canada occurred mostly first into provinces outside of Atlantic Canada [84], with federal travel measures applying to these travelers before onward travel, which may have substantially reduced the risk of disease importation to Atlantic Canada.

The cost of treating a fixed number of infections is proportionally higher in smaller economic regions, compared to larger economic regions, because of large differences in the size of the economies (see horizontal axis of Figure 4) and relatively similar costs of treating infections. The costs of strict community NPIs, such as complete business closures, might also be higher in economically larger regions relative to economically smaller regions. In Fig. 4, the pre-pandemic number of international travelers arriving to Canadian provinces and territories is shown versus their yearly Gross Domestic Product (GDP) in 2019. Areas characterised by low GDP and low travel volumes, such as Northern or Atlantic Canada, might opt for disease elimination to reduce pandemic costs by implementing travel measures to reduce the risk of community infections initiated from travel-related cases. Additionally, due to the lower travel volumes in these regions, elimination may also be more epidemiologically feasible (see point ❷). Economically larger provinces (e.g., Ontario, Quebec, British Columbia, and Alberta), might consider mitigation to be an economically preferable strategy.

**Fig. 4:**
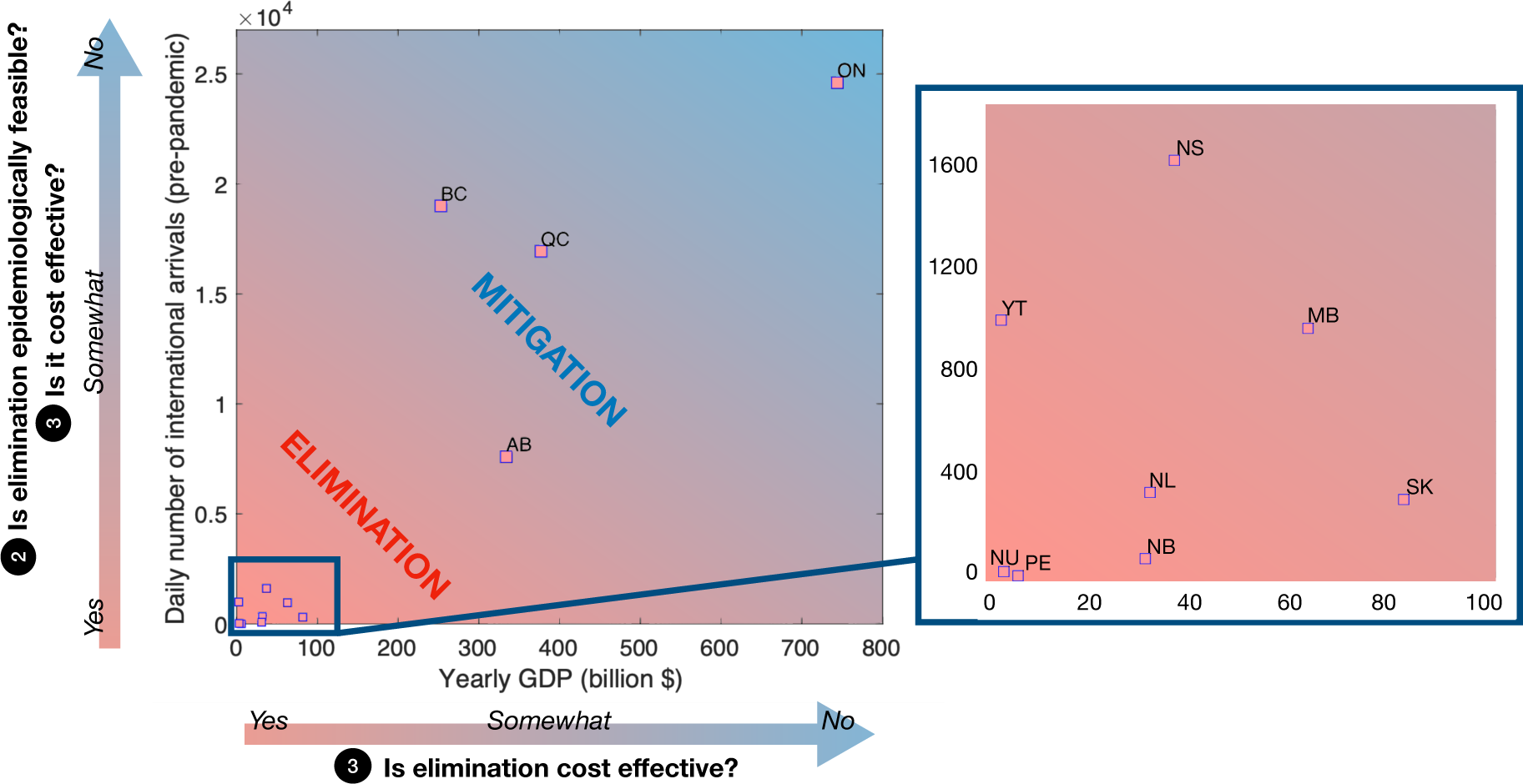
Daily number of international travelers (averaged over April 2018 to March 2019, [93]) and 2018 yearly GDP [95] obtained for different provinces and territories in Canada. Mitigation may be recommended in regions with high travel volumes, as in these regions travel measures might be less feasible and more costly. Mitigation may be recommended in regions with high travel volumes, as in these regions travel measures might be less feasible and more costly. On the other hand, limiting the number of infections through the implementation of an elimination strategy may be necessary when the cost of treating infections is high compared to the regional GDP. Note that the figure is only intended to qualitatively illustrate a possible link between arrivals, regional GDP, and costs, and we acknowledge that the nonlinearity and interdependence between these factors can make these relationships more complex. The two letter abbreviations denote Alberta (AB), British Columbia (BC), Manitoba (MB), New Brunswick (NB), Newfoundland and Labrador (NL), Northwest Territories (NT), Nova Scotia (NS), Nunavut (NU), Ontario (ON), Prince Edward Island (PE), Quebec (QC), Saskatchewan (SK), Yukon (YT). Data for the Northwest territories (NT) for the same time period are not available.

Note that the linear relationship between costs, arrivals and GDP in Fig. 4 is illustrative, and for decision-making this relationship would need to be evaluated for the specific situation. For instance, the implementation of travel measures in regions whose economies strongly depend on tourism can be very costly, especially if not well timed, and even when travel volumes are small [88, 36]. The relationship between travel volumes and community outbreak frequency may also be non-linear if efficient post-arrival travel measures (such as quarantine, or self-isolation) are in place, reducing the likelihood of community outbreaks. Finally, the relationship between the cost of treating an infection and the regional GDP indicators may not be informative when considering that a much larger number of infections may be experienced when implementing a mitigation strategy, whose relative cost in regions with higher GDP may be comparable to treating a smaller number of infections in regions with low GDP under an elimination strategy.

In addition to economic costs, social and mental health costs need to be considered when discussing NPI implementation [2]. Elimination may provide more freedoms during periods of mild restrictions, however, social interactions are substantially limited during periods of strict restrictions, which may cause high occurrence of mental health issues, such as depression and anxiety, or domestic violence [23, 25, 85]. Mitigation requires prolonged periods of moderate restrictions that can be exhausting and negatively impact the population [37, 73, 71]. Social and mental health costs of these two strategies are experienced unequally across population groups. Different social and psychological stress levels can arise depending on personal living situations, employment sector, gender, ethnicity, and social determinants of health [73], and it is misleading to report only whether elimination or mitigation guarantees lower social costs at a population level without also stratifying these costs for population groups. In many cases social costs of NPIs are to be paid in the future, and it is only recently that researchers have developed methods to determine the direct and indirect impact of NPIs implementation on a population’s health and social functioning [114, 104, 57, 28, 69]. For these reasons, it is premature to discuss which of elimination or mitigation may be preferred in terms of social cost.

## Discussion

During the pandemic, the World Health Organization recommended a risk-assessment approach that considers local epidemiology, public health measures and capacity, and contextual factors to determine if restrictions on international travel should be implemented [107]. Yet, quantitative criteria to evaluate whether an elimination or a mitigation strategy is preferable, and guidelines applicable to subnational jurisdictions, are lacking. We propose a conceptual framework to guide the decision to implement an elimination or a mitigation strategy in response to a pandemic threat. We hope that our framework will inspire new modelling approaches, for example, by expanding on the two approaches described in the supplementary information, to support and regularly reassess this decision.

So far, the focus of many epidemiological optimization models has been to determine the optimal level of social distancing needed to minimize infections, and the socio-economic costs of interventions [11, 89, 51, 38, 3, 58, 113, 4, 70, 48]. However, as noted in [38], [50] and Box 1, common modelling formulations can produce highly erroneous results when applied to situations where infection prevalence can be zero in reality. Therefore, new mathematical tools are urgently needed to quantitatively optimize the trade-off between elimination and mitigation. We also emphasize that for proper quantitative comparison of response strategies, future modelling should explicitly consider a distinction between community cases and travel-related cases, and include statistics that are relevant for the characterisation of community outbreaks occurring when an elimination strategy is implemented, such as the efficiency of border testing and quarantine policies and the probability of a traveller initiating a community outbreak [116, 44, 91, 97, 96], the expected size of such an outbreak [78, 35, 41], the probability that a community outbreak has been eliminated, such that strict NPIs might be relaxed [77, 14, 72], vaccination strategies [62, 79] and elimination exit strategies [44, 63, 103] for regions with low infection prevalence.

Our analysis supports the application of local, rather than global, public health measures and responses [66, 64, 67, 52]. Indeed, large urban areas, as compared to rural and remote areas, are characterised by dense infrastructure, greater accessibility to health care services, stronger economies, and therefore different disease dynamics and implications of public health measures. Previous studies considering the impact of travel measures to reduce the risk of SARS-CoV-2 importation have been controversial [16], and the implementation of within-state travel restrictions have been subject to legal challenges [98]. While some studies have questioned the effectiveness of travel measures [7, 24, 105], others have emphasized their importance [32, 46]. We suggest this disagreement may be due to overlooking the regional context: for example, whether community spread is occurring (Box 1), and low health care capacity, characteristics of small jurisdictions that may make elimination feasible, and the relatively low cost of implementing travel restrictions as compared to treating infections in a small economy (Fig. 2).

An elimination strategy can be implemented to maintain a healthier economy and less restrictive social distancing in economically smaller regions with low health care capacities and low travel volumes, and we present key regional and disease characteristics that can be used to assess whether elimination or mitigation may be best. Estimation of the relevant quantities (for example, by making figures similar to Fig. 3), and understanding the inter-relatedness and relative importance of key quantities (for example, by modelling the relationship between detection delay, the implementation of strict NPIs, and peak hospital occupancy, and the expected proportion of days with mild to strict restrictions, as shown in the supplementary information) is a necessary future research area to allow for a quantitative comparison of elimination and mitigation strategies for decision-making purposes.

## Data Availability

The code and data used to produce the figure are publicly available at https://github.com/ahurford/elimination-or-mitigation

## Acknowledgement

MM is grateful to the Azrieli fundation for the award of the Azrieli fellowship. MM, JA, and AH were funded by the Natural Sciences and Engineering Research Council of Canada-Public Health Agency of Canada Emerging Infectious Disease Modelling Consortium. MM and AH received funding from the Department of Health and Community Services, Newfoundland and Labrador, Canada.

