## Supplementary Information for "Is SARS-CoV-2 elimination or mitigation best? Regional and disease characteristics determine the recommended strategy"

### A. Is elimination necessary to ensure health care provision?

#### Illustrative model of peak incidence and hospital occupancy

The SIR-type model that we describe in this section can be used between the periods of elimination when a community outbreak is occurring. Regions with low testing capacity, low contact tracing capacity, or low hospital capacity may need to implement strict community NPIs as soon as community cases are detected because capacity is already close to being exceeded. Consider the dynamics of incubation, infection, and hospital occupancy:

$$\frac{dE(t)}{dt} = \beta(A(t)+I(t)) - \sigma E(t), \tag{A.1}$$

$$\frac{dA(t)}{dt} = \rho\sigma E(t) - \gamma A(t), \tag{A.2}$$

$$\frac{dI(t)}{dt} = (1-\rho)\sigma E(t) - (1-h)\gamma I(t) - h\eta I(t), \tag{A.3}$$

$$\frac{dH(t)}{dt} = h\eta I(t) - \delta H(t), \tag{A.4}$$

$$\frac{dC(t)}{dt} = (1-\rho)\sigma E(t), \tag{A.5}$$

where $E(t)$ are individuals exposed to the virus, $A(t)$ are infected asymptomatic individuals, $I(t)$ are infected symptomatic individuals (or 'active cases'), $H(t)$ are hospitalized individuals and $C(t)$ corresponds to the cumulative number of symptomatic infections. The model assumes that the fraction of individuals susceptible to infection does not change, and this quantity is understood to be included in the definition of $\beta$.

We assume that strict community NPIs are implemented at time $t = t_{\text{start}}$, and this is reflected as an instantaneous reduction in the transmission rate to $\beta = \beta_{\text{strict}}$ days$^{-1}$ at $t = t_{\text{start}}$. For an elimination strategy to be necessary it is required that

$$\beta_{\text{mild}}(A(t_0)+I(t_0)) - \gamma((1-h)I(t_0)+A(t_0)) - h\eta I(t_0) > 0, \tag{A.6}$$

such that when the disease establishes (at $t = t_0$) it spreads, and where $\beta = \beta_{\text{mild}}$ days$^{-1}$ is the transmission rate prior to the implementation of strict NPIs. For an elimination strategy to be feasible it is required that

$$\beta_{\text{strict}}(A(t_{\text{start}})+I(t_{\text{start}})) - \gamma((1-h)I(t_{\text{start}})+A(t_{\text{start}})) - h\eta I(t_{\text{start}}) < 0, \tag{A.7}$$

which is necessary for cases to decrease when strict NPIs are implemented. The variable $C(t)$ is cumulative symptomatic cases, and the daily change in $C(t)$ is incidence (also referred to as new cases; Figure A.1A, blue lines).

After the implementation of strict NPIs (at time $t_{\text{start}}$) there is a delay until active cases, $I(t)$, decline which is due to the time it takes for individuals infected just before $t_{\text{start}}$ to develop symptoms, and the average duration of this incubation period is $1/\sigma$ days (see Fig. A.1). A fraction, $\rho$, of individuals exposed do not develop symptoms, but also become infective $1/\sigma$ days after exposure. Of individuals that are active cases, a fraction $h$ will require hospitalization, and this occurs, on average, $1/\eta$ days after becoming an active case. The average duration of hospital occupancy is $1/\delta$ days, and the average duration of infectiousness for both symptomatic and asymptomatic individuals is $1/\gamma$ days.

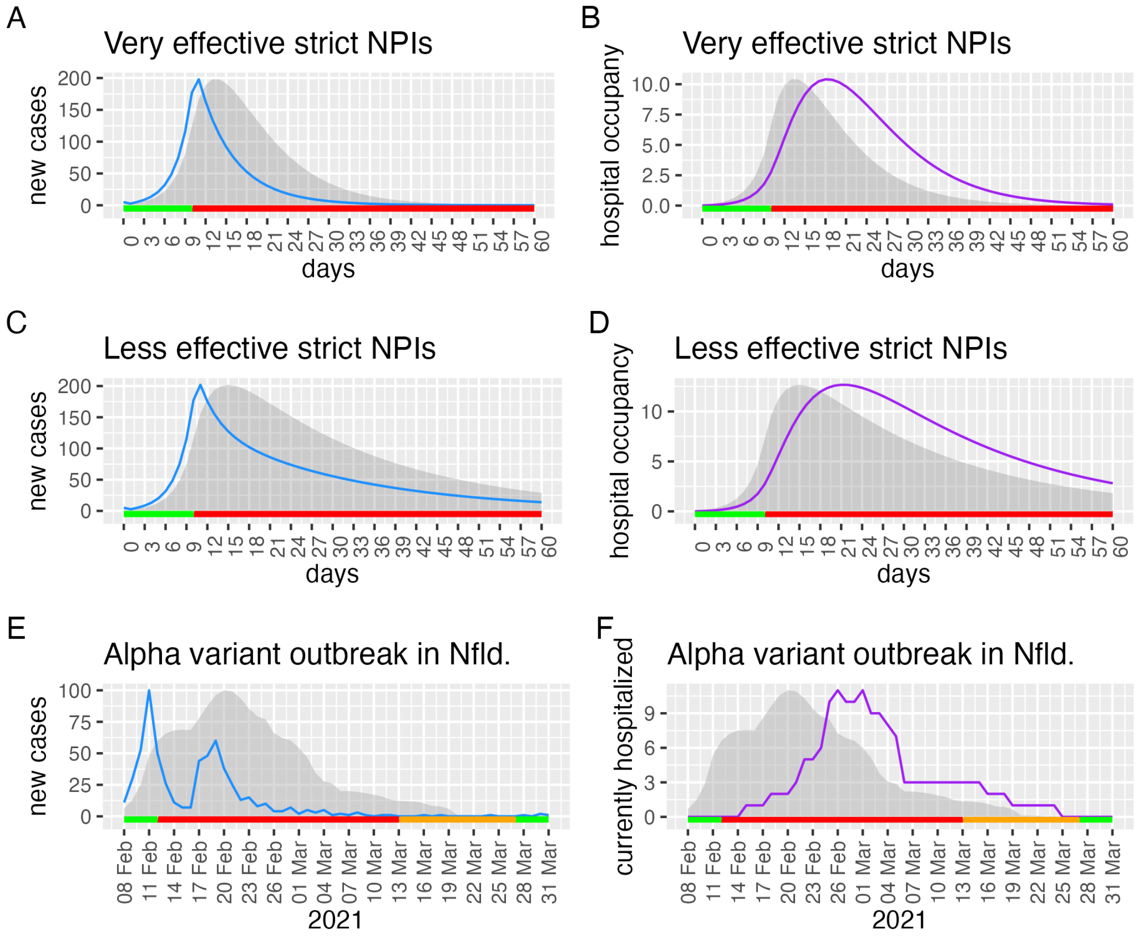


**Fig. A.1: The relationship between peak incidence, prevalence, and hospital occupancy**. (A-D) are model results (equations A.1-A.5) where strict NPIs are either very effective (top row, $\beta_{\text{strict}} = 0.01$ days$^{-1}$) or less effective (middle row, $\beta_{\text{strict}} = 0.12$ days$^{-1}$). (E-F) Data from an outbreak of the Alpha variant on the Avalon Peninsula, Newfoundland and Labrador, Canada, where strict NPIs were implemented from February 12 - March 12, 2021 (Alert level 5 on the Avalon peninsula), moderate NPIs were implemented from March 13 - March 26, 2021 (Alert level 4 on the Avalon peninsula), and mild NPIs were implemented when the whole province was at Alert level 2. In all panels (A-F), prevalence (the grey shaded area) is scaled to have the same maximum value as incidence or hospital occupancy (lines), and periods of mild, moderate, and strict NPIs are shown with green, orange, and red bars. Parameter values are: $\beta_{\text{mild}} = 2$ days$^{-1}$, $\sigma = \gamma = \delta = 1/5$ days$^{-1}$, $\rho = 0.3$, and $\eta = 1/2$ days$^{-1}$. In (A-D) strict NPIs are implemented after 10 days of mild NPIs. Initial values of the variables are: $E(t_0) = 10$, $A(t_0) = I(t_0) = C(t_0) = 5$ and $H(t_0) = 0$, with $t_0 = 0$.

Both numerical solutions to equations A.1-A.5, and reported data from the Alpha variant outbreak of SARS-CoV-2 on the Avalon peninsula of Newfoundland and Labrador in February 2021 [87], show that peak prevalence occurs a few days after peak incidence, and that peak hospital occupancy occurs a few days later (Fig. A.1). In Figure A.1, maximum prevalence is $I_{\text{max}} = 547$ (A and B), 603 (C and D), and 434 (E and F). Peak incidence (blue lines) occurs at the same time as the implementation of strict NPIs for the scenarios (A and C), and occurs one day before the implementation of strict NPIs for the Avalon peninsula outbreak (E and F). Prevalence (grey shaded region) peaks 4 days (A and B), 6 days (C and D) and 8 days (E and F) after the implementation of strict NPIs.

For the model simulations, peak hospital occupancy (purple lines) occurs 19 days (B) and 22 days (D) after the implementation of strict NPIs, and is substantially greater than hospital occupancy when strict restrictions are initially implemented. In both B and D, hospital occupancy is 2.7 when strict NPIs are implemented, while peak hospital occupancy is 10.4 (B) and 12.7 (D). For the Alpha variant outbreak on the Avalon peninsula, hospital occupancy peaks twice with 11 people hospitalized 14 and 17 days after the implementation of strict NPIs, and with 0 people hospitalized when strict restrictions were initially implemented (F).

The model (equations A.1-A.5) is designed to investigate the timing and magnitude of peak incidence and peak hospital occupancy relative to the timing of the implementation of strict NPIs. Model assumptions include that asymptomatic and symptomatic individuals are equally infectious for the same duration, symptoms and infectiousness occur concurrently, hospitalized individuals are not infectious, and there is a general lack of detail regarding contact tracing, testing, case reporting, and population heterogeneity and movement in the model. Any or all of these assumptions might be relaxed, and doing so is unlikely to change the main conclusions, for example, that peak incidence occurs at the time when strict NPIs are implemented, peak prevalence occurs a few days later, and peak hospitalization occurs a few weeks later.

In Melbourne, Australia, frequently peak incidence did not coincide with the implementation of strict NPIs [51, 103]. In considering Melbourne, Australia, there may be a violation of our requirement that elimination is feasible (Eq. A.7) as defined above. The code to produce Fig. A.1 is available at `https://github.com/ahurford/elimination-or-mitigation`.

### B. Frequency of community outbreaks and disease severity for different COVID-19 variants of concern

#### B.1 Estimating the proportion of days with mild restrictions when implementing an elimination strategy

When an elimination approach to SARS-CoV-2 is applied, strict restrictions are implemented to bring the number of cases back to zero whenever a community outbreak occurs, and restrictions are relaxed afterwards. We are interested in broadly estimating the expected percentage of days when mild restrictions are implemented for different viral variants (i.e., the percentage of 'green' zones during the time period characterised by $T_i$ in Fig. 1).

We know that the SARS-CoV-2 Alpha variant was 1.5 more transmissible than the original variant [62], that the Delta variant is 1.6 more transmissible than Alpha [124], and that the Omicron variant is 3.3 more transmissible than Delta [76]. The Alpha variant arrived in Canada at the end of December 2020 [109]. Newfoundland and Labrador experienced a community outbreak of Alpha variant in February 2021, and no successive outbreak until the end of April 2021 [27]. Therefore, we use 120 days as an estimate for the expected time between community outbreaks ($T_i$) due to the Alpha variant. Respective estimates for other variants are provided in Table B1.

Additionally, we consider an elimination strategy that is implemented when $I_{\text{max}}$ active cases are already present in the community, and we calculate the time needed to reduce the number of active cases from $I_{\text{max}}$ to $I_{\text{end}}$. This time interval is shown as $T_e$ in Fig. 1, where $I_{\text{max}}$ and $I_{\text{end}}$ represent, respectively, the number of active cases when the outbreak is detected and when strict restrictions are released. Variation in the number of active cases over time is based on the infection transmission rate $\beta$ and recovery rate $\gamma$, and can be formulated as:

$$\frac{dI(t)}{dt} = I(t)(\beta - \gamma), \tag{A.8}$$

with $\beta/\gamma = R_c$ being the control reproductive number. We base our calculation on the assumption that once the outbreak is detected, the number of cases decays exponentially over time, where $I(t) = I_{\text{max}} e^{\gamma(R_c-1)t}$ and $R_c < 1$ when strict measures are implemented. We also assume that no other outbreak occurs during this time, and we do not account for travel-related cases that do not cause community cases. We find that

$$I_{\text{end}} = I_{\text{start}} e^{\gamma(R_c-1)T_e} = I_{\text{end}} \quad \Longleftrightarrow \quad T_e = \frac{\ln(I_{\text{start}}/I_{\text{end}})}{\gamma(1-R_c)}. \tag{A.9}$$

We consider that the control reproduction number with strict restrictions implemented is $R_c = 0.1$ for the original strain, $I_{\text{max}} = 50$ and $I_{\text{end}} = 1$ and calculate the corresponding control reproduction number for different variants of concern taking into account their increased transmissibility [62, 124, 76]. We consider a recovery rate of 1/7 days$^{-1}$. Finally, we calculate the expected percentage of days for when mild restrictions can be implemented, given an elimination strategy, as

$$\text{\% of days with mild restrictions} = \begin{cases} \dfrac{T_i - T_e}{T_i} \times 100 & \text{for } T_i > T_e \\ 0 & \text{otherwise} \end{cases} \tag{A.10}$$

Estimates are provided in Table B1. Results are shown in Fig. 3.

#### B.2 Estimating disease severity

Using the estimates of hospitalization rates for different viral variants in vaccinated and unvaccinated individuals [82], and Newfoundland and Labrador vaccination rates [28], we estimate the average number of hospitalizations per 1000 infections in Newfoundland and Labrador when variants were established in the province [109]. Estimates are provided in Table B1 and Fig. 3.

**Table B1:** Estimates of SARS-CoV-2 variants epidemiological characteristics in Newfoundland and Labrador (NL) used to produce Fig. 3.

| | **Original** | **Alpha** | **Delta** | **Omicron** |
|---|---|---|---|---|
| Data of establishment in NL [7] | Mar 14, 2020 | Feb 12, 2021 | Apr 28, 2021 | Dec 15, 2021 |
| Percent of NL population vaccinated with one dose when the variant was established [6] | – | 0.93% | 26.46% | 6.41% |
| Percent of NL population vaccinated with two doses when the variant was established [6] | – | 0.78% | 1.86% | 85.33% |
| Average probability of hospitalization for unvaccinated individuals [5] | 2.3% | 3.84% | 5.50% | 1.82% |
| Average probability of hospitalization for individuals vaccinated with one dose [5] | – | 1.79% | 2.14% | 1.27% |
| Average probability of hospitalization for individuals vaccinated with two doses [5] | – | – | 1.73% | 1.01% |
| Average hospitalizations per 1000 cases, given NL vaccination rates when the variant was established | 23 | 38 | 34 | 11 |
| Estimated time between community outbreaks (parameter $T_i$) | 180 days | 120 days | 75 days | 23 days |
| Estimated duration of periods of strict restrictions to bring the number of cases from 50 to 1 (i.e, parameter $T_e$ in Eq. (A.9)) | 30 days | 32 days | 36 days | 132 days |
| If elimination is implemented, estimated % of days during which mild restriction are implemented | 83.1% | 73.2% | 52.0% | 0% |
